# Continued T12 transmission and shared antibiotic resistance during 2018-2023 *Vibrio cholerae* outbreaks in Cameroon

**DOI:** 10.1101/2024.08.26.24303680

**Authors:** Sen Claudine Henriette Ngomtcho, Blaise Mboringong Akenji, Roland Ndip, Andrew S Azman, Yanick Carolle Tayimetha, Etienne Guenou, Sylvain Engamba, Marie Claire Assoumou Okomo, Justin Lessler, Shirlee Wohl

**Author notes:** Correspondence (S.W.), (S.C.H.N.).

## Abstract

Seventh pandemic *Vibrio cholerae* was first identified in Cameroon in 1971, causing several sporadic disease clusters with few cases. More recent years have seen larger cholera outbreaks, but the mechanism behind these periodic outbreaks is poorly understood, and it is unclear the degree to which antibiotic resistant strains contribute to disease burden and spread. We used whole genome sequencing to characterize 14 *V. cholerae* isolates from the 2020 and 2021-2023 cholera epidemics in Cameroon. All these isolates belonged to the T12 lineage, and most showed the same antimicrobial resistance (AMR) pattern regardless of year. This suggests that cholera outbreaks in Cameroon are, at least in part, a continuation of the outbreaks previously reported in 2018 and as far back as 2012. This finding has important implications for cholera management since it suggests the ongoing presence of pathogenic cholera even in years with few reported cases. Similarly, the AMR results suggest the need for new treatment approaches, as resistance to many common antibiotics was found even within our limited sample set. As such, whole genome sequencing should be implemented in low-income countries such as Cameroon to improve disease surveillance and to detect and predict pathogen antibiotic resistance profiles.

## INTRODUCTION

Since the first documented cholera pandemic in 1817, seven *Vibrio cholerae* pandemics have been recorded [1]. The seventh pandemic originated in Southeast Asia in 1961 and is still ongoing. It reached the African continent in 1970, first affecting West African countries (Guinea-Bissau and Guinea Conakry), and then reaching Central Africa, and specifically Cameroon, in 1971 [2, 3]. Despite having been cholera-free before 1970, Africa now bears the major burden of the global cholera cases, with over 4 million cholera cases and 143,000 deaths reported in Africa per year [2].

Cameroon reported its first cholera cases in February 1971. This was followed by a 20-year period characterized by sporadic disease clusters with few cases. Following this period, larger outbreaks were reported in 1991, 1996, 1998, 2004, 2010, 2011, and over 46,000 suspected cholera cases and 1,800 deaths were reported between 2004 and 2013 [4]. Minimal cases were reported in intervening years, and the mechanism behind these periodic cholera outbreaks remains poorly understood. Specifically, it is unclear if successive outbreaks are caused by sustained circulation of existing strains, or if each is caused by a new introduction.

More recent cholera outbreaks in Cameroon have followed a similar trend, despite increased efforts to improve surveillance nationwide. On May 18, 2018, two suspected cases of cholera were reported in the North Region of Cameroon and ultimately confirmed as *Vibrio cholerae* by bacterial culture. Cases associated with this outbreak spread into the Far North Region and continued until December 2019, resulting in a total of 1212 suspected cases and a 5.1% case fatality rate (CFR) in the North Region and 350 cases (CFR: 4.6%) in the Far-North [5].

After a brief period of few cases, the Ministry of Health in Cameroon declared another outbreak following laboratory confirmation of *Vibrio cholerae* in late January 2020. 18 health districts were affected in four neighboring administrative regions (South-West, Littoral, South and Center) and the outbreak lasted throughout the final weeks of 2020, causing 1892 cases (CFR: 4.2%) [6]. After several months of minimal cases (<200 in total), a new outbreak was declared in Cameroon on October 29, 2021. As of October 2023, over 21,000 suspected cases have been reported (CFR: 2.4%) (**Supplementary Data 1**) [7]. This 2023 epidemic has spread to the six main regions of the country, with the central Littoral Region becoming the epicenter of the epidemic. Understanding how each of these outbreaks spread and the relationship between them may inform approaches to cholera containment in Cameroon and have implications for outbreak management elsewhere.

Molecular characterization of seventh pandemic *V. cholerae* may be one important tool for understanding cholera outbreak emergence and spread. Already, genomic data has led to new insights about global cholera transmission and has highlighted the important role of transmission within and between Asia and Africa [8, 9]. Combining genomic and epidemiological data has allowed scientists to better understand the dynamics of ongoing outbreaks, and advances in sequencing technology have recently made whole genome sequencing more feasible even in low-resource settings. Previous studies initially identified three waves of seventh pandemic *V. cholerae* [10] which were later broken down into at least 14 plausible introduction events from Asia into Africa, termed T1–T14 [8, 9, 11]. These lineages can be used to identify the approximate source of case clusters; for example, cases caused by the same lineage as a previous outbreak might suggest undetected sustained transmission, while identification of a new lineage could suggest that new cases were caused by a new international introduction.

Even so, identification of *V. cholerae* strains responsible for cholera in Cameroon epidemics has been largely limited to bacteriological culture, without more advanced molecular analyses like whole genome sequencing [12]. To better understand the cholera dynamics in Cameroon and Sub-Saharan Africa as a whole, we characterized *V. cholerae* O1 isolates from the 2018-2019, 2020, and 2021-2023 (still ongoing) epidemics, with the goal of identifying and comparing the *V. cholerae* lineages in circulation in Cameroon, and ultimately using this information to better understand cholera transmission dynamics in order to prioritize public health action. As antimicrobial resistance is becoming an increasingly serious threat to public health [13], we also determined the antimicrobial resistance profile of these samples, in the hopes of providing information that can be used to limit the spread of pathogenic drug-resistant *V. cholerae*.

## RESULTS

To better understand *V. cholerae* transmission patterns in Cameroon, we sequenced 55 isolates from epidemiological and laboratory-confirmed cases from the 2018, 2020 and 2021 epidemics (**Table 1**). We selected these isolates to ensure representation as many regions of Cameroon as possible (samples were sequenced from eight of the ten regions of Cameroon, see **Figure 1**). We sequenced these isolates at the National Public Health Laboratory in Cameroon using the Oxford Nanopore platform and generated whole genome sequences (>97% reference genome coverage) from 14 of the 55 isolates (25% success rate) (**Supplementary Data 1**). Although we successfully obtained sequences from six included regions, none of the isolates from the 2018 epidemic achieved our 97% coverage threshold and were therefore excluded from phylogenetic analysis. We observed higher sequencing success rate from samples from the most recent epidemics, perhaps due to contamination issues associated with poor long-term storage conditions of older samples.

**Table 1.** Isolates selected for sequencing and antimicrobial resistance phenotyping.

| Year | Isolates Available | Isolates Sequenced | High quality Genomes | Disc Diffusion Results | Sequenced + Disc Diffusion | High quality + Disc Diffusion |
| --- | --- | --- | --- | --- | --- | --- |
| 2019 | 29 | 2 | 0 | 2 | 2 | 0 |
| 2020 | 30 | 20 | 4 | 9 | 9 | 3 |
| 2022 | 28 | 22 | 9 | 10 | 10 | 5 |
| 2023 | 43 | 11 | 1 | 33 | 0 | 0 |
| **TOTAL** | **130** | **55** | **14** | **54** | **21** | **8** |

**Figure 1.**
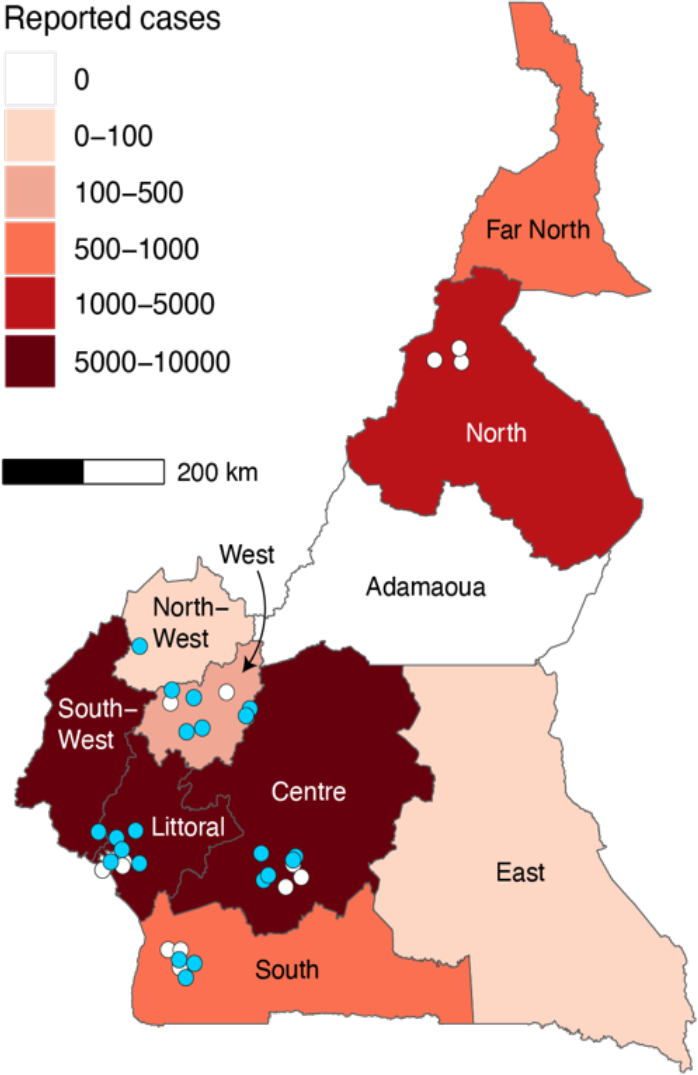
Spatial distribution of *V. cholerae* isolates selected for sequencing. Blue: high quality genomes included in downstream analyses; white: sequenced isolates that did not produce high quality genomes (see also **Supplementary Data 2**). The number of cholera cases reported between 18 May 2018 – 17 October 2023 are shown in shades of red.

We generated a maximum likelihood tree using the 14 successful sequences to better understand how *V. cholerae* O1 was circulating in Cameroon over time and space (**Figure 2**). Combining these sequences with previously published global *V. cholerae* genomes demonstrated that they belonged to the T12 lineage [9], which was shown to be circulating in Cameroon as recently as 2018 [12] and is the only lineage to have been observed in Cameroon since 2009 (**Figure 2**, see also **Supplementary Data 3**). Sequences from both 2020 and 2022 are found in this lineage, with no evidence for sequences belonging to other lineages.

**Figure 2.**
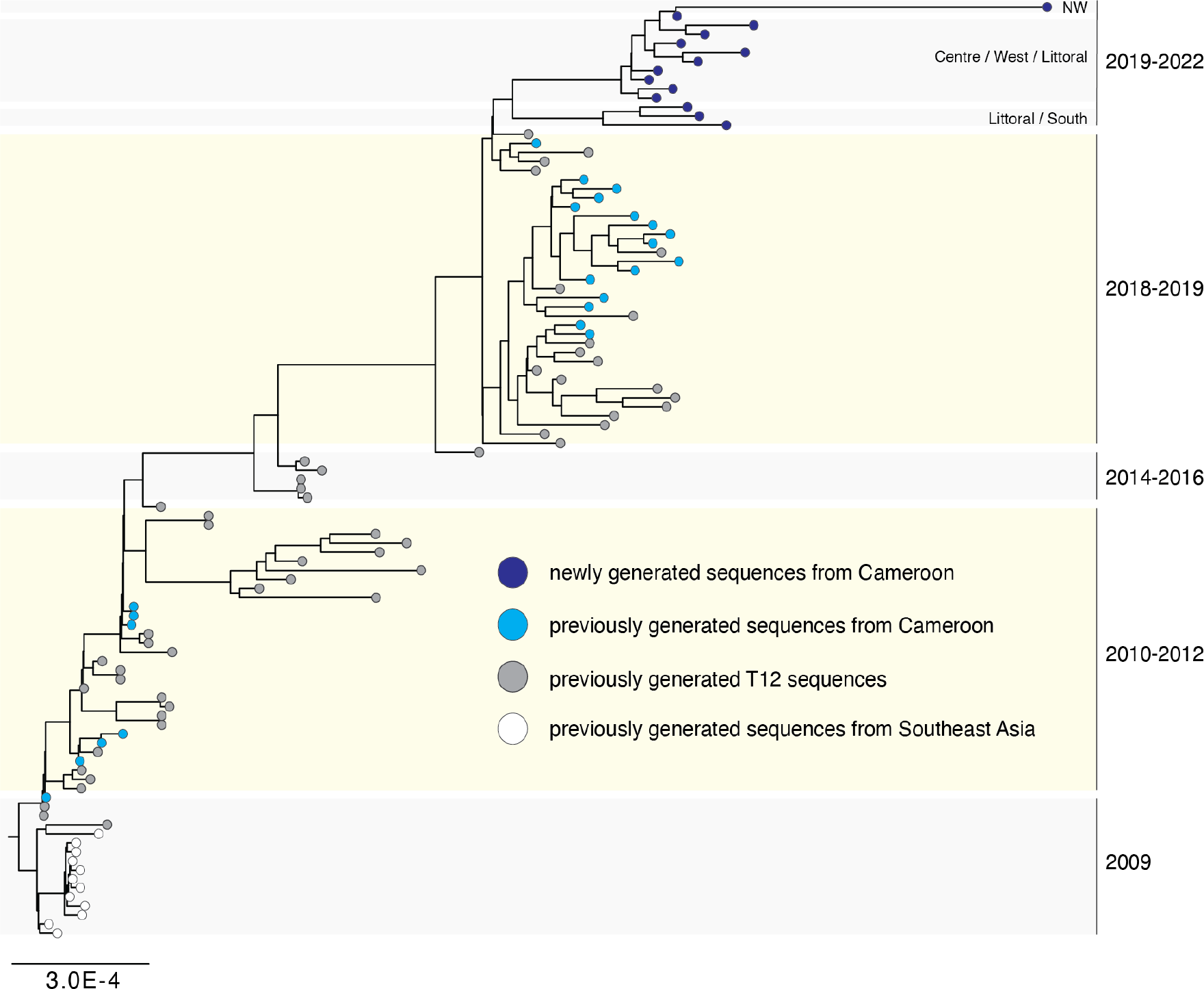
Maximum likelihood tree of the *V. cholerae* T12 lineage. Sequences generated in this study cluster together and are shown in dark blue. Previously generated sequences from Cameroon are shown in light blue, and other previously generated sequences from the T12 lineage are shown in gray. Groups of newly generated sequences are labeled by the region in Cameroon the corresponding isolates were collected in (NW = North West). Scale bar: nucleotide substitutions per site.

Combined with the results presented in Ekeng et al., our observation of T12 sequences in both the 2020 and 2021 outbreaks provides strong evidence for continuous circulation of this lineage within Cameroon. From our data, it is clear that the T12 lineage is even more widespread than previously thought, as we observed this lineage in three new epidemiological areas: the North West, West, and South Regions (previously published sequences from the 2018 outbreak were exclusively from the North, Littoral and Center Regions of the country)

(**Figure 2**). The new sequences generated in this study form two distinct Cameroon-specific sub-clusters that both contain 2020 and 2022 sequences—as well as sequences from multiple regions of the country— suggesting circulation and evolution within Cameroon during the 2020 and 2021 outbreaks. These sub-clusters appear to be descendants of the West Africa T12 sub-cluster from 2018, supporting prior evidence for transmission between the neighboring countries of Nigeria, Niger, and Cameroon.

To understand sequenced isolates that did not generate a high-quality *V. cholerae* O1 genome, we looked more closely at samples that generated a large number of reads that did not map to the N16961 *V. cholerae* O1 reference sequence. We identified 24 samples that generated at least 39,000 reads (the minimum number of reads used to assemble a high-quality *V. cholerae* O1 genome in samples sequenced as part of this study) that were ultimately excluded from our analysis. Of these, more than half were completely missing the ctxA gene, according to the typing results (**Supplementary Data 1**). Although poor sequencing quality may still explain these results, the existence of potential non-O1 *V. cholerae* among these isolates should not be excluded.

We also explored the results of antimicrobial resistance (AMR) testing to better understand the relationship between isolates. We performed AMR testing on 54 isolates (including 13 of the 14 that produced *V. cholerae* O1 whole genome sequences) using disk-diffusion methods (**Supplementary Data 4**). We found that resistance to most of the antibiotics tested was mixed (except for erythromycin, to which nearly all tested isolates were resistant), and that these results did not cluster by time or place of sample collection. This finding highlights why doxycycline, tetracycline, and ciprofloxacin are some of the primary antibiotics used to combat cholera disease in Cameroon, though approximately 50% of isolates tested in this study were resistant to doxycycline and/or tetracycline (55% and 50%, respectively), suggesting a need for continued monitoring of AMR, including the extent to which AMR phenotype may correlate with genetic signatures.

## DISCUSSION

Our finding of the T12 lineage in both 2020 and 2022 in Cameroon supports previous evidence suggesting continuous circulation of the lineage in the country (or at least in the region) since it was first observed in 2009. This suggests that cholera containment efforts should, at least in part, focus on reducing community transmission, either by reducing transmission routes between linked regions, or focusing on prevention measures such as drinking and using clean water, washing hands with soap more often, and using adequate sanitation in regions with high case numbers.

One challenge to cholera containment may be the fact that *V. cholerae* strains circulating in Africa have reportedly been expanding the number of antibiotic classes to which they confer resistance [8]. This is cause for concern, as these antibiotic drugs have long been used for treating severe cholera cases. Our results show the presence of at least some resistant isolates in all drugs tested, highlighting the need for continued monitoring of antimicrobial resistance, even when cases seem to be derived from the same persistent lineage.

Despite the clear implications of our genomic findings for containment of the T12 lineage, genomic surveillance in low-resource public health settings continues to be challenging, in part because samples are not always collected and preserved with sequencing in mind. For example, we found that less than 30% of *V. cholerae* isolates sequenced in this study produced a high-quality genome. Because we performed only reference-based assemblies to the *V. cholerae* O1 reference, it is possible that some of the failed samples were because they were of a different serogroup (see typing results in **Supplementary Data 1**). However, most failed sequences are likely due to low input DNA concentrations due to poor storage conditions, inappropriate transport conditions from the field to NPHL, or lack of resources (i.e., transport media, technology support) needed for storage, transport, or sequencing. Notably, almost all samples from 2023 failed because of lack of sequencing platform support. Investment in surveillance systems that include routine whole genome sequencing may address some of these sequences, especially those caused by poor long-term storage conditions.

Since the introduction of *V. cholerae* into Africa from Asia in 1961, a limited number of isolates from central and western sub-region of Africa have been sequenced, with fewer than 100 (as of time of writing) sequences available from Cameroon, most of which were sequenced abroad. In the present study, we greatly increase the fraction of sequences generated in-country, which may ultimately decrease the time between sample collection and generation of actionable genomic results. This highlights the necessity for public health authorities in low-income countries to further develop genomic surveillance strategies of pathogens with epidemic potential such as cholera.

## MATERIAL AND METHODS

### Data Availability

Raw data for all sequenced isolates is available under NCBI BioProject accession: PRJNA954566. Source data files have been provided for all figures as Supplementary Data 1-4. Laboratory protocol and bioinformatics pipeline used for analysis are publicly available at https://github.com/HopkinsIDD/minion-vc.

### Ethics Statement

Samples used in this study were collected as part of ongoing outbreak investigations for the monitoring of cholera in Cameroon. Only leftover clinical diagnostic samples (already stored as bacterial isolates) were used for sequencing. No identifying patient information was included in this study, and only bacterial isolates were sequenced (i.e., no chance of sequencing human data). Ethical approval was granted by the University of Buea (Cameroon) Faculty of Health Sciences Institutional Review Board (2023/1974-02/UB/SG/IRB/FHS).

### Sample selection, collection, and cholera confirmation

In this study, we selected randomly from all *V. cholerae* isolates stored at NPHL from the 2018-2023 outbreaks (see **Table 1**). We excluded samples for which we did not obtain *V. cholerae* growth after replating the isolate on TCBS and Muller Hinton media.

These isolates were originally collected and cultured as follows: isolates were generated from fresh (within 24-48 hours of symptom onset) stool samples from suspected cholera cases during the three most recent cholera outbreaks in Cameroon (starting in 2018, 2020, and 2021). These samples were initially collected in sterile screw-capped stool recipients by laboratory technicians and trained community healthcare workers in communities and health facilities in districts that notified suspected cases. Specifically, two sterile cotton swabs were immersed into the stool and aseptically inserted into a screw tube containing 10 ml Cary-Blair transport medium. For children who could not produce stools on demand, samples came from rectal swabs collected using moist cotton swabs and dipped into Cary-Blair transport media. Samples were placed in a triple package insulated box and transported to the National Public Health Laboratory (NPHL) in Cameroon within a maximum of 48 hours.

At NPHL, each stool sample collected during the outbreaks was first directly inoculated on thiosulfate citrate bile salt sucrose (TCBS) and incubated at 37°C for 24 hours. Immediately after culturing the first TCBS dish, sample was introduced into alkaline peptone water (APW) and incubated at 37°C for 8 hours. A second TCBS dish was then inoculated with the stool sample enriched on APW and incubated at 37°C. After 24 hours of incubation, characteristic *V. cholerae* colonies were selected and finally subcultured on Muller Hinton agar and incubated at 37°C for 24 hours. Colonies resulting from this culture on Muller Hinton medium were resuspended in 5ml of sterile saline solution to obtain a 0.5 McFarland inoculum and used for the phenotypic identification of *V. cholerae* based on biochemical characteristics and serotyping. Biochemical identification was done using API 20E biochemical galleries (BioMerieux, USA). Each well was inoculated using a sterile pipette and then incubated for 16-24 hours at 37°C and the reactions were noted as either positive or negative. Serotyping was done by agglutination tests using commercial *V. cholerae* Ogawa and Inaba antisera kits (Deben Diagnostics, India) according to the manufacturer instructions.

### Antibiotic Susceptibility Testing

Antibiotic susceptibility testing was performed by disk-diffusion method [15] for the following antimicrobial drugs: amoxicillin (20μg), amoxicillin/clavulanic acid (10-20μg), cephalotin (20 μg), cefotaxim (30μg), chloramphenicol (30μg), gentamicin (15μg), nalidixic acid (20μg), doxyciclin (30μg), tetracycline (30μg), polymixin B, azythromicin (15μg) and ciprofloxacin (30μg). The tested antibiotics were chosen based on the European Committee on Antimicrobial Susceptibility Testing (EUCAST, http://www.eucast.org). These antibiotics and the agar swabbing inoculation technique described below underwent quality control using *Escherichia coli* ATCC 25922 reference strains before use for *V. cholerae*. In brief, after 5 min of incubation of *V. cholerae* inoculum on Mueller Hinton agar at room temperature, antibiotic discs were applied on the medium. Plates were incubated at 37°C overnight. Interpretation was done by measuring inhibition diameters around the disks using a caliper. Sensitivity, resistance, or the intermediate phenotypes of the strain to antibiotics was interpreted applying the appropriate EUCAST guidelines. Abricate software (www.github.com/tseemann/abricate) and the Comprehensive Antibiotic Resistance Database (CARD) (www.card.mcmaster.ca/analyze/rgi) online tool were used to identify genes and single-nucleotide polymorphisms (SNPs) associated with antibiotic resistance.

### DNA extraction and quantification

DNA extraction was performed at NPHL on *V. cholerae* isolates from the 2018 and 2020 outbreaks. Samples stored at -80°C in glycerol-brain heart infusion medium were allowed to thaw at room temperature, and then were re-plated on Mueller Hinton agar (MH) and TCBS. Samples from the 2021 outbreak were directly plated on TCBS culture medium. All plates were incubated at 37°C overnight. 2 to 4 colonies were then inoculated into 4ml of sterile Bacto™Peptone (alkaline peptone water) and incubated at 37°C overnight. A shaker was used during this incubation. Afterwards 1ml of cloudy culture media was used for DNA extraction using the Qiagen QIAmp DNA Mini Kit, with a final elution volume of 150 μL. The extracted DNA concentration was directly measured using a Quantus Fluorometer, following the instructions of the manufacturer.

### Oxford Nanopore library construction and sequencing

Library preparation and sequencing was performed at NPHL in Yaoundé, Cameroon. We used modified version of the Oxford Nanopore 1D Native barcoding genomic DNA (with EXP-NBD104, EXP-NBD114, and SQK-LSK109) as described in Ekeng et al. [12]. The extracted DNA was diluted to 1 μg in 49 μL with nuclease-free water and 48μL of this solution was used for library preparation. For samples with a DNA concentration below 20 ng/μL, 48μl of extracted DNA was used regardless of the final amount (<1μg). Sequences were prepared in 6 different batches (**Supplementary Data 1**).

After library preparation following the Oxford Nanopore protocol, we diluted the final library to a final volume of 12 μL in Elution Buffer and combined with 37.5 μL Sequencing Buffer and 25.5 μL Loading Beads. This mixture was loaded onto a primed R9.4.1 FLO-MIN106 Oxford Nanopore MinION flow cell. For each sequencing library, the MinION was run for 48 hours.

### Reference-based genome assembly

As described in Ekeng et al. [12], sequencing runs were basecalled using Guppy version 4.0.4 with the flip-flop model (dna_r9.4.1_450bps_fast.cfg). Porechop (https://github.com/rrwick/Porechop) was used for demultiplexing and adapter removal, and we filtered out low quality sequencing reads using Filtlong (https://github.com/rrwick/Filtlong) with the following options: ‘filtlong --keep_percent 90 --target_bases 800000000’.

Using the resulting filtered FASTQ files, we performed reference-based assembly following the bioinformatics pipeline available here: https://github.com/HopkinsIDD/minion-vc. We used the N16961 strain (Genbank accession: AE003852/AE003853) as a reference for these seventh pandemic O1 sequences and required coverage of at least 100x to call a base at each site across the genome. Assemblies with less than 97% coverage of the reference genome and a median read depth (across the complete N16961 reference genome) below 100 were excluded from subsequent phylogenetic analysis.

### *Vibrio cholerae* typing

Typing was performed by mapping reads from sequenced isolate to reference genes ctxA, tcpA_classical, tcpA_eltor, toxR, wbeO1, and wbfO139, with Genbank accession numbers AF463401, M33514.1, KP187623.1, KF498634.1, KC152957 and AB012956, respectively [16]. We mapped filtered reads to each reference using minimap2 version 2.17 [17] using scripts included as part of the publicly available bioinformatics pipeline noted above. Using samtools, we counted the number of reads mapping to each gene reference, the fraction of the gene covered by mapped reads, and the depth of coverage of each site across the reference gene of interest.

### Maximum likelihood estimation

We concatenated reference-based assemblies generated during this study to 1323 previously published *V. cholerae* O1 whole genome sequences [8–10, 12, 18, 19] also assembled to the N16961 reference. These background sequences were selected to represent most of the publicly available whole genome sequences (**Supplementary Data 3**). We masked recombinant sites in these sequences as previously described, first with a selection of fixed masked sites from Weill et al. 2017 [8] (available at: https://figshare.com/s/d6c1c6f02eac0c9c871e) and then using Gubbins version 3.2.1 [20]. To generate maximum likelihood trees, we ran IQ-TREE version 1.4.4 [21] using the ModelFinder [22] option, which identified TVM+F+ASC+G4 as the best nucleotide substitution model, and 1000 ultrafast bootstrap replicates [23]. The final tree was rooted on the taxa labeled 5174_7_5 (accession: ERR025382), as described in Weill et al. 2019 [9] and Ekeng et al. 2021 [12]. We visualized the resulting tree in FigTree version 1.4.4 (http://tree.bio.ed.ac.uk/software/figtree/).

